# The effect of smoking on COVID-19 symptom severity: Systematic review and meta-analysis

**DOI:** 10.1101/2020.08.15.20102699

**Authors:** Askin Gülsen, Burcu Arpinar Yigitbas, Berat Uslu, Daniel Drömann, Oguz Kilinc

## Abstract

**Background:** Coronavirus disease 2019 (COVID-19) is caused by severe acute respiratory syndrome coronavirus 2 (SAR2-COV-2), and was first identified in Wuhan, China in December of 2019, but quickly spread to the rest of the world, causing a pandemic. While some studies have found no link between smoking status and severe COVID-19, others demonstrated a significant one. The present study aimed to determine the relationship between smoking and clinical COVID-19 severity via a systematic meta-analysis approach.

**Methods:** We searched the Google Scholar, PubMed, Scopus, Web of Science, and Embase databases to identify clinical studies suitable for inclusion in this meta-analysis. Studies reporting smoking status and comparing non-severe and severe patients were included. Non-severe cases were described as mild, common type, non-intensive care unit (ICU) treatment, survivors, and severe cases as critical, need for ICU, refractory, and non-survivors.

**Results:** A total of 16 articles detailing 11322 COVID-19 patients were included. Our meta-analysis revealed a relationship between a history of smoking and severe COVID-19 cases (OR=2.17; 95% CI: 1.37–3.46; *P* <.001). Additionally, we found an association between the current smoking status and severe COVID-19 (OR=1.51; 95% CI: 1.12–2.05; *P* <.008). In 10.7% (978/9067) of non-smokers, COVID-19 was severe, while in active smokers, severe COVID-19 occurred in 21.2% (65/305) of cases.

**Conclusion:** Active smoking and a history of smoking are clearly associated with severe COVID-19. The SARS-COV-2 epidemic should serve as an impetus for patients and those at risk to maintain good health practices and discontinue smoking.

## Introduction

Coronavirus disease 2019 (COVID-19), which is caused by severe acute respiratory syndrome coronavirus 2 (SAR2-COV-2), was first identified in Wuhan, China in December of 2019. It has subsequently spread across the world, causing a global pandemic. This highly contagious disease has thus far infected 5.2 million people worldwide and killed approximately 337000 patients, yielding a case fatality rate (CFR) that varies between 0.7 and 12.7% (average: 6.4%).^[1,2]^

COVID-19 primarily targets lung epithelial cells, causing viral pneumonia and acute respiratory distress syndrome (ARDS), especially in elderly patients. Therefore, mortality is higher in the elderly and in patients with at least one accompanying comorbid disease.^[3]^ In the last report issued by the Centers for Disease Control and Prevention Institute, the incidence of respiratory disease was 9.2% in patients diagnosed with a severe COVID-19 clinical course.^[4]^ Chronic obstructive pulmonary disease (COPD) and asthma are also common comorbidities in severe cases and are reported in 10.8% and 17.0%, respectively, of hospitalized patients aged ≥18 years with COViD-19.^[5]^

However, it has been reported that COVID-19 progresses more severely in COPD patients.^[6]^ Given that smoking plays an important role in the etiopathogenesis of COPD, it may have a similar effect on symptoms. In a recent meta-analysis of smoking and COVID-19 severity, smoking was found to not increase the severity of COVID-19 (odds ratio [OR], 1.69; 95% Cl: 0.41-6.92). However, only five studies were included in this meta-analysis, and heterogeneity among the studies was low (I^2^ = 38%).^[7]^

In another meta-analysis involving more studies, COVID-19 severity was found to increase with smoking (OR, 1.97; 95% Cl: 0.95-4.10).^[6]^ To increase the statistical power of this analysis, case reports and series were also included, not just case-control studies. As previously, the heterogeneity of the studies included here was also low (I^2^=44%).^[6]^ The different results of both meta-analyzes create confusion about the issue of smoking status. Given diverging findings in the existing literature, we systematically reviewed English-language studies to investigate whether smoking was associated with a more severe clinical course of COVID-19.

## Material and Methods

All guidelines listed in the Preferred Reporting Items for Systematic Reviews and Meta Analyses (PRISMA) statement were followed in conducting this meta-analysis.^[8]^ Our methodological systematic review and meta-analysis of pooled case-control studies were recorded in the International Prospective Register of Systematic Reviews (https://www.crd.york.ac.uk/prospero; registration number: CRD42020180173).

### Study search strategy

We searched the Google Scholar, PubMed, Scopus, Web of Science, and Embase databases to identify clinical studies suitable for inclusion here. Only English-language articles published between December 2019 and April 15, 2020 were included. Case series/reports, commentaries, reviews, and editorial letters were excluded. Unpublished articles were not searched, but “ahead of print” and “first only” articles with a DOI link in the given database were included. The following search terms were used: “COVID-19,” OR “SARS-COV-2,” OR “novel coronavirus (nCOV),” AND “smoking,” OR “tobacco,” AND “clinical features,” OR “characteristic,” AND “severity,” OR “severe”.

### Inclusion and exclusion criteria

Inclusion criteria were as follows: (i) studies that examined COVID-19 patients older than 18 years and diagnosed COVID-19 according to WHO criteria; (ii) observational, cross-sectional, prospective, or retrospective studies; (iii) studies which compared smokers with mild and severe COVID-19; (iv) studies comparing smokers with survivors and non-survivors of COVID-19; and (v) studies comparing smokers who developed COVID-19 that required intensive-care unit (ICU) and non-ICU treatment.

Exclusion criteria were as follow: (i) studies from the same medical center that did not contribute to meta-analysis, (ii) non-English-language studies; (iii) case reports/series; (iv) reviews; (v) editorial letters; (vi) studies lacking a full-text (unavailable or not yet published); (vii) studies without a DOI; and (viii) studies with small sample sizes (<50 patients) because of low statistical power.

### Data extraction

A researcher (AG) scanned study titles and abstracts obtained via an initial database search and included relevant ones in a secondary pool. Next, two independent researchers (AG and BU) evaluated the full text of these articles to determine whether they met study inclusion criteria. Any disputes were resolved by discussion and negotiation with a third investigator (BAY). Agreed-upon studies were included in the final meta-analysis.

The following data were obtained from all studies: title, first author, publication year, location, sample size, age (median), history of smoking (former or current), and distribution of non-severe or severe COVID-19 status. Given these groups, we were able to compare non-severe (mild, common type, non-ICU, survivors) and severe (critical, severe, ICU, nonsurvivors) COVID-19 cases.

### Risk of bias (quality) assessment

Two researchers (BAY and BU) evaluated the collected articles using the Newcastle-Ottawa scale (NOS) for study quality and risk of bias.^[9]^ This scale evaluates three aspects of each study: (i) selection (0 to 4 points), (ii) comparability (0 to 2 points), (iii) and detection of the outcome of interest (0 to 3 points). According to obtained total score, study quality was rated as follows: 0-3 points (poor), 4-6 points (intermediate), and 7-9 points (high). Studies with a high risk of bias were excluded during the meta-analysis phase.

### Statistical analyses

OpenMeta Analyst version 10.10 (https://www.cebm.brown.edu/open_meta) was used to calculate odds ratios with 95% confidence intervals, which are depicted using forest plots. Quantitative numbers were measured in terms of total numbers and percentages (n, %). The odds-ratio of smokers among non-severe COVID-19 and severe COVID-19 cases, as well as survivors and non-survivors, were calculated. Heterogeneity was evaluated with Cochran’s Q, χ^2^, and the Higgins I^2^ test. The Higgins I^2^ test uses a fixed effects model when I^2^ <50%, and a random effects model when I^2^ >50%. When heterogeneity was detected, a sensitivity adjustment was made to determine its source. This procedure was performed by leaving one study out of the analysis at a time, with a fixed effects model used after excluding heterogeneity. Publication bias was evaluated with Statsdirect version 3.2.10 (StatsDirect Ltd, Cambridge, UK) and visualized using Begg’s funnel plots and Egger’s test. P-values (twosided) were considered statistically significant if *P* <.05.

## Results

Our initial search of international databases using the keywords described above yielded 265 articles. After duplicate articles were excluded, 197 articles remained. After article titles and abstracts were evaluated for appropriateness, a final 34 articles were found to meet study inclusion criteria. In addition, 18 of the studies for which full texts were obtained were excluded because they met exclusion criteria. A total of 16 articles met all our criteria. The relevant PRISMA study flow chart is shown in Figure-1. A quality analysis of the pooled articles included in this meta-analysis was done by applying NOS. As a result, overall quality was found to be moderate (NOS score min: 5, max: 8). Results of a quality analysis of the pooled articles are available in table 1.

**Figure 1.**
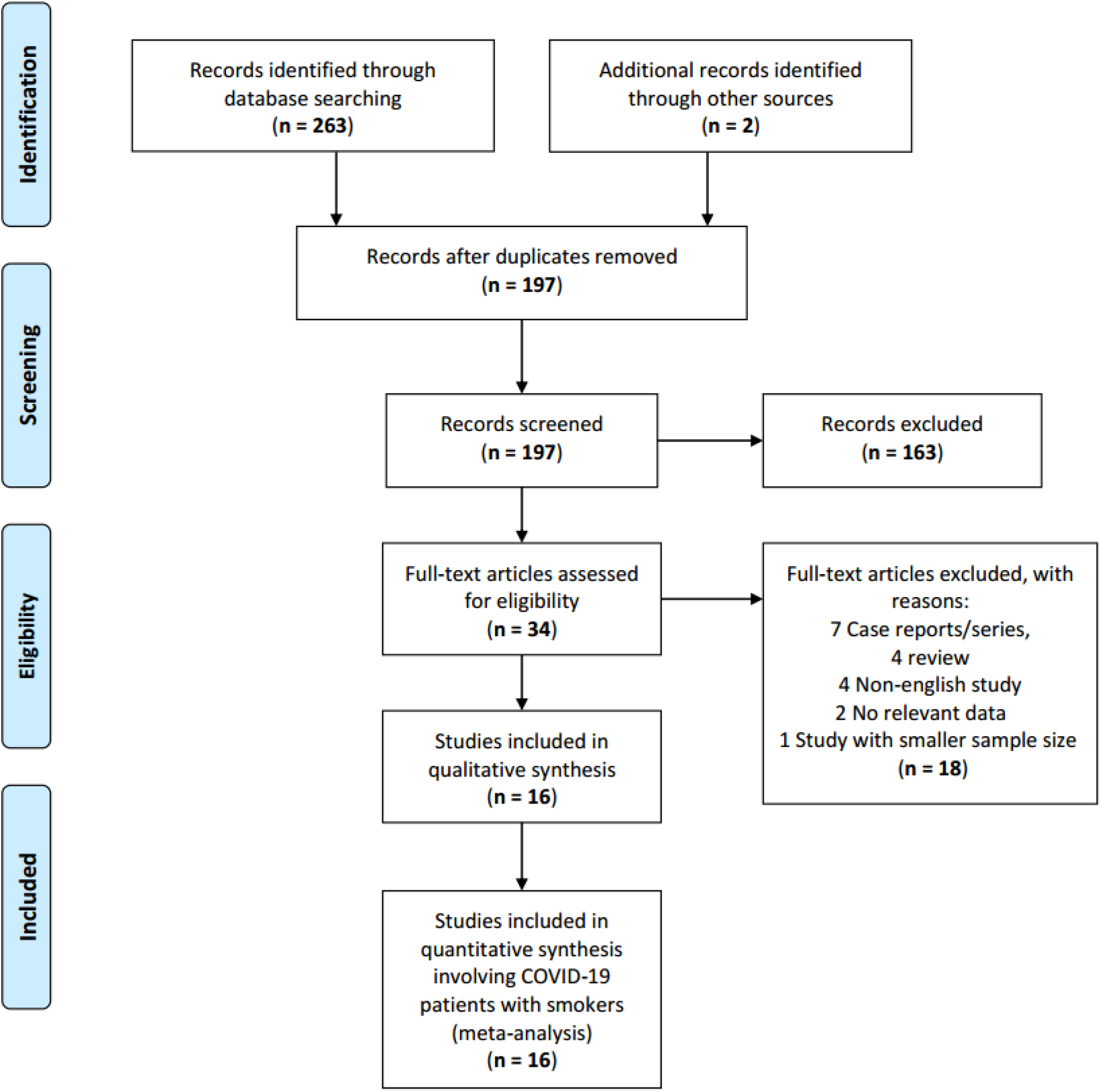
PRISMA flow-diagram of study selection procedures.

**Table-1.**
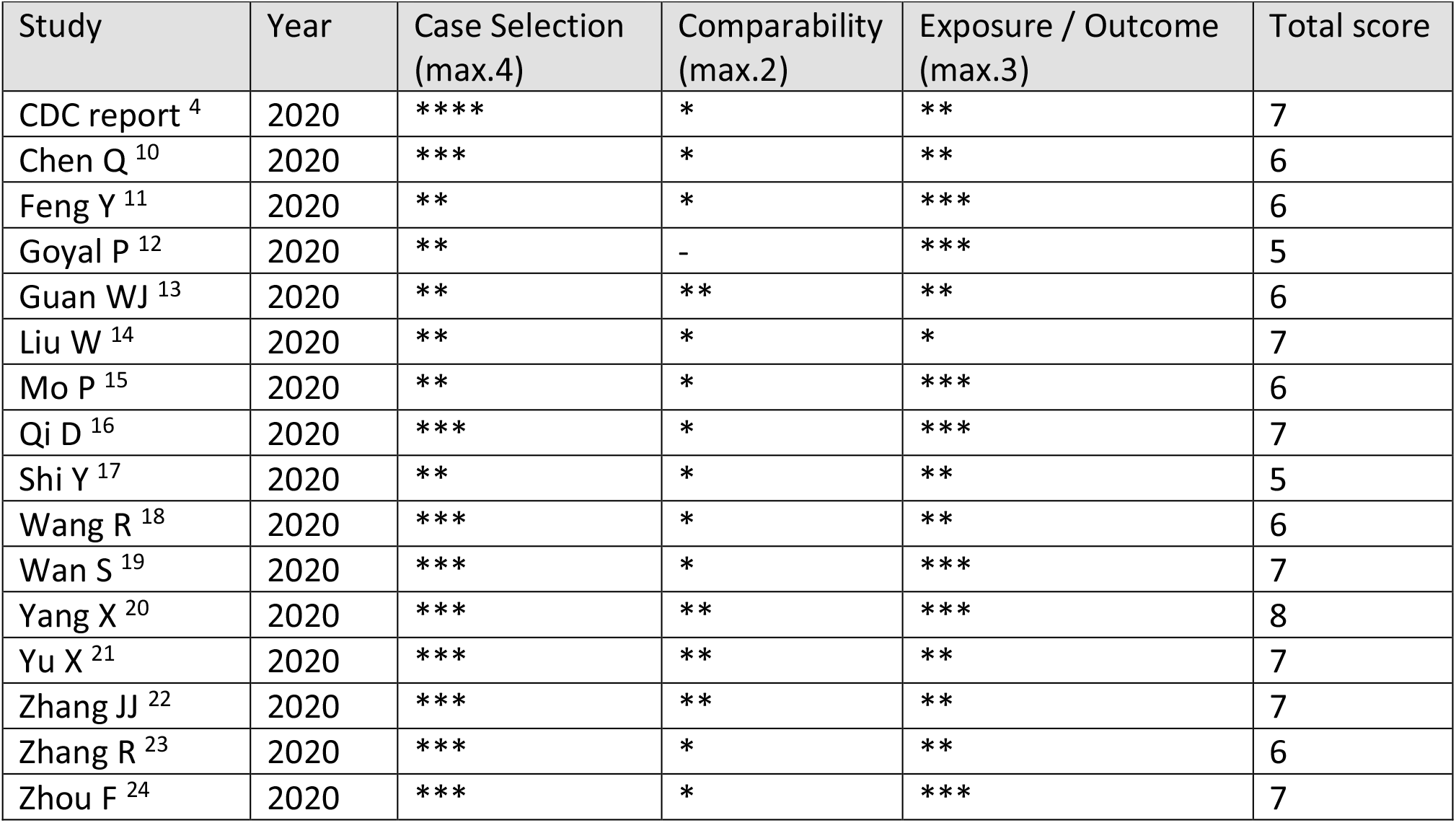
Newcastle-Ottawa scale for quality assessment and risk of bias

### Features of the included studies

In the initial meta-analysis pool, 11322 patients with COVID-19 from 16 studies (4, 10-24) were included. However, given CDC reporting guidelines (4), 525 patients whose hospitalization statuses were reported as “unknown” were excluded from the final analysis, yielding a final 10797 patients. Of these, 9414 (87.2%) were non-severe COVID-19 patients, while 1383 (12.8%) were severe or critical COVID-19 patients who required intensive care. Among the severe or critical patients, smoking prevalence ranged from 3.6-19.9% (average: 8.4%). While nine studies provided current smoker information, three provided both current smoker and former smoker information and seven provided only historical smoking information. All of the included studies were retrospective (six multicenter and 10 single-center). A summary of the studies included in this meta-analysis is available in table 2.

**Table-2.** Features of the studies included in the meta-analysis

| First Author | Location of Patients | Study Design | Comparison | n | Age, year | Prevalence of smoking | Former smokers | Current smokers |
| --- | --- | --- | --- | --- | --- | --- | --- | --- |
| CDC report <sup>4</sup> | - | R, MC | Non-ICU vs ICU | 7162 | - | 261 (3.6) | 165 (2.3) | 96 (1.3) |
| Chen Q <sup>10</sup> | Taizhou, China | R, O, SC | Non-severe vs severe | 145 | 47.5 | 15 (10.3) | - | - |
| Feng Y <sup>11</sup> | Wuhan, Anhui, Shanghai | R, O, MC | Moderate vs Severe, Critical | 454 | 53.0 | 44 (9.7) | - | - |
| Goyal P <sup>12</sup> | Newyork, USA | R, MC | Non-IMV vs IMV | 393 | 62.2 | 20 (5.1) | - | 20 (5.1) |
| Guan WJ <sup>13</sup> | 552 hospitals in China | R, MC | Non-severe vs severe | 1085 | 47.0 | 158 (12.6) | 21 (1.9) | 137 (12.6) |
| Liu W <sup>14</sup> | Wuhan, China | R, SC | Stabilization vs Progression | 78 | 38.0 | 5 (6.4) | - | - |
| Mo P <sup>15</sup> | Wuhan, China | R, SC | General vs refractory | 155 | 54.0 | 6 (3.9) | - | 6 (3.9) |
| Qi D <sup>16</sup> | Chongqing, China | R, MC | Non-severe vs Severe | 267 | 48.0 | 53 (19.9) | - | - |
| Shi Y <sup>17</sup> | Zhejiang, China | R | Mild vs severe | 487 | 46.0 | 40 (8.2) | - | - |
| Wang R <sup>18</sup> | Fuyang, China | R, SC | Non-critical vs critical | 125 | 49.4 | 16 (12.8) | - | 16 (12.8) |
| Wan S <sup>19</sup> | Chongqing, China | R, SC | Mild vs severe | 135 | 47.0 | 9 (6.7) | - | 9 (6.7) |
| Yang X <sup>20</sup> | Wuhan, China | R, O, SC | Survivors vs non-survivors | 52 | 51.9 | 2 (10.0) | - | - |
| Yu X <sup>21</sup> | Shanghai, China | R, SC | Non-severe vs severe | 333 | 50.0 | 26 (7.8) | - | - |
| Zhang JJ <sup>22</sup> | Wuhan, China | R, SC | Non-severe vs severe | 140 | 57.0 | 9 (6.4) | 7 (5.0) | 2 (1.4) |
| Zhang R <sup>23</sup> | Wuhan, China | R, SC | Common typ vs severe | 120 | 45.4 | 6 (5.0) | - | 6 (5.0) |
| Zhou F <sup>24</sup> | Wuhan, China | R, O, MC | Non-survivor vs survivor | 191 | 56.0 | 11 (6.0) | - | 11 (6.0) |

Data are presented as "n" or "n (%)". Age-related data are given as "median year". n, population; R, retrospective; CS, cross-sectional, O, observational; IMV, invasive mechanic ventilation; ICU, intensive care unit; SC, single-center; MC, multi-center.

### History of smoking and COVID-19 severity

The prevalence of smoking among patients with mild to moderate COVID-19 was 493/9414 (5.2%), while among severe and critical cases it was 173/1383 (12.5%). A meta-analysis revealed a significant relationship between smoking history and severe COVID-19 (random effect model, OR=2.17; 95% CI:1.37–3.46; *P* <.001) (Table-3). A funnel plot was used to evaluate publication bias and revealed substantial heterogeneity between the pooled studies (I^2^=71.4%; *P* <.001) (Figure 2b). A sensitivity analysis conducted after removing two studies (Qi D et al.^[16]^ and Zhang R et al.^[23]^), which were primary causes of heterogeneity, revealed that smoking was significantly related to COVID-19 severity (fixed effect model, OR=1.85; 95% CI:1.50–2.29; *P* <.001) (Figure 2c). Furthermore, this updated analysis had low heterogeneity (I^2^=40.6%; p=.06), while a funnel pilot and Egger’s test revealed no publication bias (tau^2^=-0.87, *P* =.16) (Figure 2d).

**Table-3.**
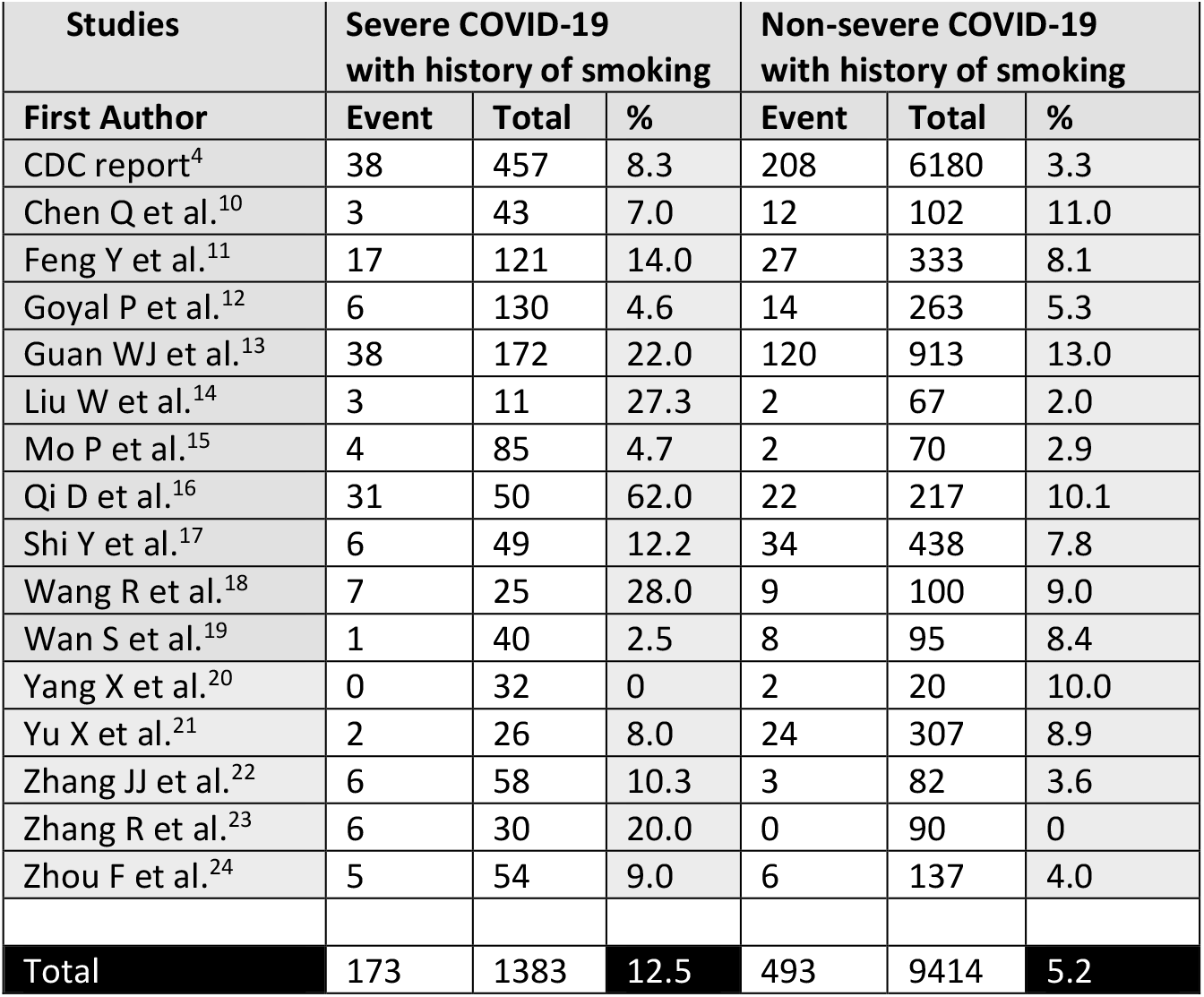
The relationship between history of smoking* and the severity of COVID-19

| Studies | Severe COVID-19 with history of smoking |  |  | Non-severe COVID-19 with history of smoking |  |  |
| --- | --- | --- | --- | --- | --- | --- |
|  | Event | Total | % | Event | Total | % |
| CDC report<sup>4</sup> | 38 | 457 | 8.3 | 208 | 6180 | 3.3 |
| Chen Q et al.<sup>10</sup> | 3 | 43 | 7.0 | 12 | 102 | 11.0 |
| Feng Y et al.<sup>11</sup> | 17 | 121 | 14.0 | 27 | 333 | 8.1 |
| Goyal P et al.<sup>12</sup> | 6 | 130 | 4.6 | 14 | 263 | 5.3 |
| Guan WJ et al.<sup>13</sup> | 38 | 172 | 22.0 | 120 | 913 | 13.0 |
| Liu W et al.<sup>14</sup> | 3 | 11 | 27.3 | 2 | 67 | 2.0 |
| Mo P et al.<sup>15</sup> | 4 | 85 | 4.7 | 2 | 70 | 2.9 |
| Qi D et al.<sup>16</sup> | 31 | 50 | 62.0 | 22 | 217 | 10.1 |
| Shi Y et al.<sup>17</sup> | 6 | 49 | 12.2 | 34 | 438 | 7.8 |
| Wang R et al.<sup>18</sup> | 7 | 25 | 28.0 | 9 | 100 | 9.0 |
| Wan S et al.<sup>19</sup> | 1 | 40 | 2.5 | 8 | 95 | 8.4 |
| Yang X et al.<sup>20</sup> | 0 | 32 | 0 | 2 | 20 | 10.0 |
| Yu X et al.<sup>21</sup> | 2 | 26 | 8.0 | 24 | 307 | 8.9 |
| Zhang JJ et al.<sup>22</sup> | 6 | 58 | 10.3 | 3 | 82 | 3.6 |
| Zhang R et al.<sup>23</sup> | 6 | 30 | 20.0 | 0 | 90 | 0 |
| Zhou F et al.<sup>24</sup> | 5 | 54 | 9.0 | 6 | 137 | 4.0 |
| **Total** | **173** | **1383** | **12.5** | **493** | **9414** | **5.2** |

**Figure 2.** (A), Prevalence of smoking in severe COVID-19 patients; (B), Funnel plot for meta-analysis of the prevalence of smoking in severe COVID-19 patients; (C), Prevalence of smoking among severe COVID-19 patients after the exclusion of studies by Qi D et al.^16^ and Zhang R et al.^23^; (D), Funnel plot for the meta-analysis of smoking among severe COVID-19 patients after the exclusion of studies by Qi D et al.^16^ and Zhang R et al.^23^.

### Smoking status and COVID-19 severity

Only 10 of the pooled studies provide information about current smoking, specifically (n=305). Current smoking prevalence in patients with mild to moderate COVID-19 was 229/7850 (2.9%), while in severe or critical cases it was 65/1134 (5.7%). However, in 10.7% (978/9067) of non-smokers, COVID-19 disease was severe, while in active smokers it was severe in 21.2% (65/305) of cases (Table-4). This clearly demonstrates a significant relationship between current smoking and severe COVID-19 patients (fixed effect model, OR=1.51; 95% CI:1.12-2.05; z=2.65; *P* =.008), with an approximate 1.5-fold increased risk of ICU admission, symptom, severity and mortality in smokers (Figure 3a). In this analysis, we used a fixed effect model due to the low level of heterogeneity (I^2^=48.9%; *P* =.04). A funnel pilot and Egger’s test revealed no publication bias (tau^2^=0.22, *P* =.77) (Figure 3b). A sensitivity analysis conducted after removing one study (Zhang R et al.^[23]^) revealed a similar relationship with severe COVID-19 (fixed effect model; OR=1.36; 95% CI:1.00-1.87; p=0.05) and low heterogeneity (I^2^:34.0%; *P* =.14) (Figure 3c). A funnel pilot and Egger’s test further revealed no publication bias (tau^2^=-0.31, *P* =.67) (Figure 3d).

**Table-4.**
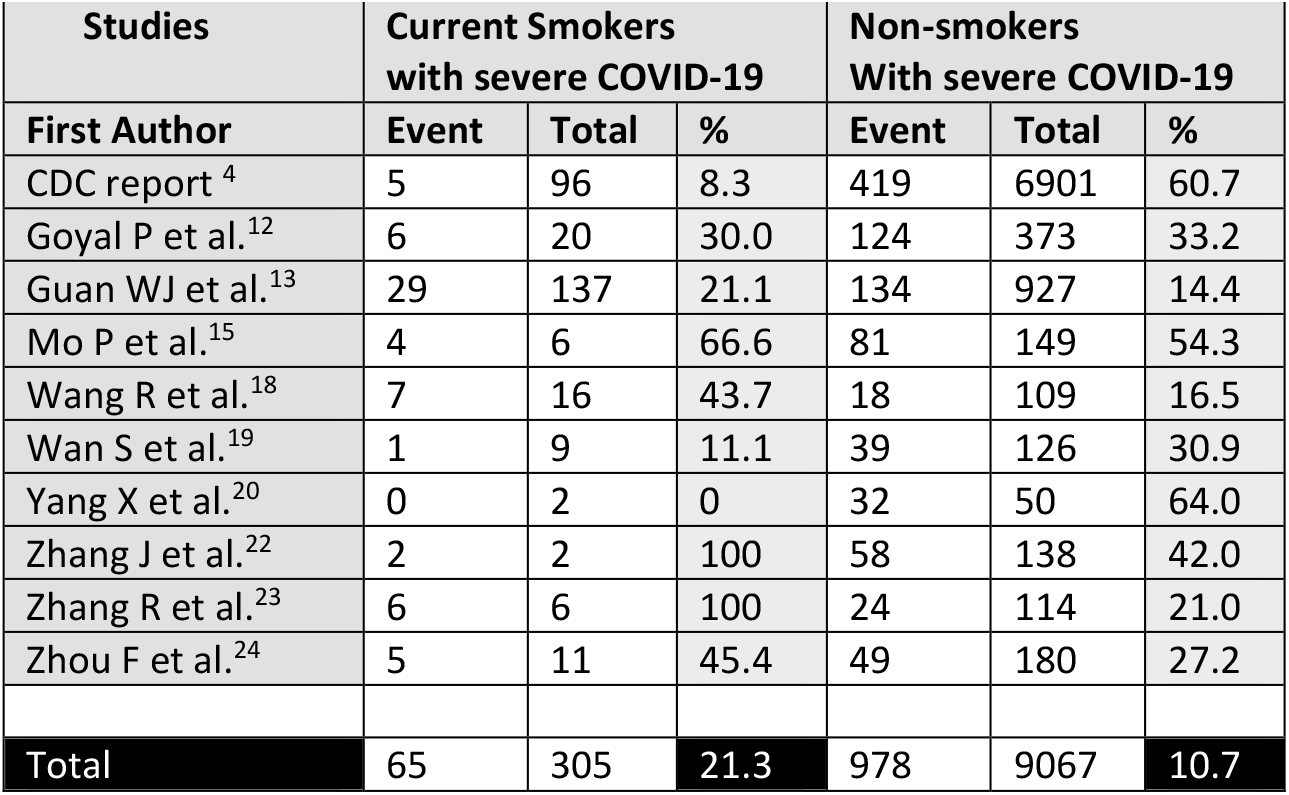
The relationship between smoking status and the severe COVID-19

| Studies | Current Smokers with severe COVID-19 |  |  | Non-smokers With severe COVID-19 |  |  |
| --- | --- | --- | --- | --- | --- | --- |
|  | Event | Total | % | Event | Total | % |
| CDC report <sup>4</sup> | 5 | 96 | 8.3 | 419 | 6901 | 60.7 |
| Goyal P et al.<sup>12</sup> | 6 | 20 | 30.0 | 124 | 373 | 33.2 |
| Guan WJ et al.<sup>13</sup> | 29 | 137 | 21.1 | 134 | 927 | 14.4 |
| Mo P et al.<sup>15</sup> | 4 | 6 | 66.6 | 81 | 149 | 54.3 |
| Wang R et al.<sup>18</sup> | 7 | 16 | 43.7 | 18 | 109 | 16.5 |
| Wan S et al.<sup>19</sup> | 1 | 9 | 11.1 | 39 | 126 | 30.9 |
| Yang X et al.<sup>20</sup> | 0 | 2 | 0 | 32 | 50 | 64.0 |
| Zhang J et al.<sup>22</sup> | 2 | 2 | 100 | 58 | 138 | 42.0 |
| Zhang R et al.<sup>23</sup> | 6 | 6 | 100 | 24 | 114 | 21.0 |
| Zhou F et al.<sup>24</sup> | 5 | 11 | 45.4 | 49 | 180 | 27.2 |
| **Total** | **65** | **305** | **21.3** | **978** | **9067** | **10.7** |

**Figure 3 (A)**, Comparison of smoking status in severe COVID-19 patients; (B),Funnel plot for the meta-analysis of smoking among severe COVID-19 patients; (C), Comparison of smoking status among severe COVID-19 patients after the exclusion of a study by Zhang R et al.^23^; (D), Funnel plot for the meta-analysis of smoking among severe COVID-19 patients after the exclusion of a study by Zhang R et al.^23^.

## Discussion

The present meta-analysis contained 16 studies and revealed that those with a history of smoking and active smokers had significantly increased risk for severe COVID-19. Previous studies have reported a similar result.^[14,18,23]^ In addition, one prior meta-analysis reported that disease severity increased given a history of smoking (OR: 1.98).^[6]^ Our analysis adds to this growing consensus in the literature, invoking more studies and examining both a history of smoking and current smoking status in risk for increased COVID-19 severity.

Five prior studies have reported results which disagree with those presented here—namely, no significant difference between patients with and without a smoking history in terms of COVID-19 severity.^[10,15,21,22,24]^ In addition, a meta-analyses performed by Lippi et al.^[7]^ failed to find a relationship between active smoking and severe COVID-19 on Chinese patients, and another meta-analysis indicates that active smoking is not a predisposing factor for hospitalization.^[25]^ First of all, both meta-analysis^[7,25]^ was not systematic, and included no assessment of publication bias or study quality. As such, it should be considered only preliminary. Hence, the present systematic meta-analysis offers a more detailed view as it covers 16 studies. The heterogeneity was low to moderate and, after sensitivity adjustments, the prevalence of a history of smoking was found to be 5.2% in non-severe COVID-19 cases and 12.5% in severe cases. Furthermore, the prevalence of active smoking was 2.9% in non-severe COVID-19 cases and 5.9% in severe cases. Therefore, severe COVID-19 was observed almost 1.5 to 2 times more frequently in history of smoking and current smoking groups. There are studies^[4,11,14,16,18,23]^ and meta-analysis^[6,26-34]^ results supporting this finding.

In addition, a recent meta-analysis revealed that active smokers are at higher risk of mortality and serious complications.^[27]^ Interestingly, this meta-analysis indicated that serious complications were observed in 48% of former smokers and 24% of current smokers during COVID-19 course.^[27]^ This appears to be because former smokers have longer exposure times or accompanying diseases such as COPD due to smoking. This issue was explained recently in a rapid non-systematic meta-analysis conducted by Guo FR, which showed that COPD increased the development of severe COVID-19 by 4.38 times.^[28]^ These findings are supported by another rapid non-systematic meta-analysis, suggesting that smoking increases severity and mortality of COVID-19 in hospitalized patients.^[29]^ In addition, history of smoking has been reported to increase the progression of COVID-19 disease.^[30]^ There is growing evidence to support WHO’s statements that ’smokers are at a higher risk of developing severe COVID-19 and consequent death.^[31]^ It seems very clear that the pandemic period is an opportunity to quit smoking due to the possibility of encountering worse clinical outcomes and complications in patients with smoking history.^[32]^

The relationship between smoking and illness severity due to other respiratory viruses has been investigated previously. In one recent study, COPD patients who smoked had decreased leukemia inhibitory factor levels during respiratory syncytial viral infection, particularly during exacerbations, which may have contributed to lung tissue injury.^[35]^ In addition, smoking may reduce the protective effect of influenza vaccines in elderly patients and increase risk of hospitalization.^[36]^ As a result, adverse effects on the lungs in smokers may aggravate the symptom severity of viral infections.

Furthermore, novel SARS-CoV-2 uses angiotensin converting enzyme (ACE)-2 in the lungs to enter cells and cause infection. In patients using ACE inhibitors or Angiotensin II receptor blockers (ARBs) for hypertension, upregulated ACE2 expression impacts on disease course remains unclear. However, the Council on Hypertension of the European Society of Cardiology does not recommend discontinuation of ACEI and ARB medications for all patients.^[37]^ The ACE-2 upregulation may increase infectiousness and therefore infection risk, as the SARS-COV-2 virus uses this receptor for host entry. Paradoxically, it is stated that it may be useful in protecting people from acute lung injury.^[35]^

In one recent study, ACE-2 gene expression was up-regulated in the airway epithelium of COPD patients and active smokers, thereby suggesting a mechanism by which risk for severe COVID-19 increases in smokers.^[39]^ This upregulation may occur through the α7 subtype of nicotine acetylcholine receptors (α7-nAChR) in people who smoke and consequently consume nicotine.^[40]^ Therefore, α7-nAChR antagonists (e.g., α-conotoxin, methyllycaconitine), which may be used to assist with smoking cessation, should be investigated for their potential impact on ACE-2 expression and thus SARS-COV-2 entry into the host.^[41,42]^ Furthermore, α7-nAChR and ACE-2 may be potential therapeutic targets, a topic which requires further investigation.

Common comorbidities including cardiovascular disease, chronic kidney disease, diabetes, and hypertension are associated with increased risk for severe COVID-19.^[43]^ Similarly, COPD is also associated with a poor clinical progression and poor outcomes despite treatment, and it was reported that the risk of severe COVID-19 was observed 4 times higher in patients with pre-existing COPD than in patients without COPD.^[6]^ This may be explained by systemic and chronic inflammation, diminished respiratory function and capacity, and COPD-related respiratory failure in some patients. The prevalence of COPD was previously reported to be 2.8% in non-smokers, 7.6% in former smokers, and 15.2% in active smokers.^[44]^ Given this, smoking and COPD comorbidity should be considered together as a single risk factor for severe COVID-19.

Another important point for consideration is that acute respiratory distress syndrome and cytokine storms, which are seen in some severe COVID-19 patients, are key causes of mortality in COVID-19. In these patients, pro-inflammatory cytokines such as interleukin (IL)-1, IL-2, IL-6, IL-8, IL-17, interferon-γ, and TNF-α are elevated, which affect patients clinical symptoms and severity.^[45]^ Nicotine is a cholinergic agonist that acts via the α7-nAChR pathway, and also inhibits pro-inflammatory cytokines such as IL-1, IL-6, and TNF, but has no effect on IL-10, an anti-inflammatory cytokine.^[46]^ Therefore, nicotine may be beneficial for the treatment of severe COVID-19 when delivered via chewing gum, inhaler/aerosol, or patch.^[47,48]^ This potential therapeutic application should be clarified with future clinical research, however.

Several factors limit interpretation of the present study. First, the vast majority of studies included here were retrospective epidemiological studies conducted in China. Second, some included studies did not distinguish between former or current smoking status. Third, studies classified COVID-19 cases broadly as follows: (i) mild to moderate: mild, non-severe, common type, did not require ICU care, and COVID-19 survivors and (ii) severe: severe, critical, required ICU care, and non-survivors. Given these limitations, caution should be exercised while interpreting our results. Future studies may respond to these issues by defining disease severity more clearly and by obtaining more detailed information about smoking habit.

In addition to classical tobacco smoking behavior, water pipe and electronic cigarette use should not be overlooked, as these modes of consumption may increase contamination risk due to repetitive hand interactions with the mouth, carrying cigarette packets in the pockets, and blowing of smoke. Similarly, exposure to passive smoke can alter ACE-2 gene expression and cause immune system changes. Naturally, these patients also have the potential to have severe COVID-19 symptoms, and smoke exposure is a potential risk factor for those around the patient, including their friends and family.

Future studies should continue to collect nicotine consumption information, including the number of cigarettes smoked per day, passive exposure, and degree of COPD, and should evaluate the dynamics of interactions between cigarette smoking and COVID-19. In addition, the effects of various smoking habits (e.g., mild versus heavy consumption, water pipe use, and electronic cigarettes use) on the transmission of SARS-COV-2, the clinical severity of COVID-19, and the clinical progression of COVID-19 should be investigated. Finally, the relationship between COPD severity and COVID-19, and the potential therapeutic effect of nicotine on severe COVID patients should also be examined and clarified. In addition, clinicians can pay more attention to the history of smoking of COVID-19 patients, and more further research may aim to determine mechanisms that drive or decrease this risk.

## Conclusion

The present meta-analysis revealed that active smoking and a history of smoking are significantly associated with increased COVID-19 symptom severity. The SARS-COV-2 epidemic should serve as an impetus for patients and those at risk to maintain good health practices and discontinue smoking.

## Data Availability

The data [data.oma and data.xls] used to support the findings of this study are available from the corresponding author upon request.

## Acknowledgements

Funding of the Federal Ministry for Science and Education (BMBF), German Center for Lung Research, is gratefully acknowledged.

